# How the prospect of a clinical trial impacts decision-making for predictive genetic testing in amyotrophic lateral sclerosis

**DOI:** 10.1101/2024.09.30.24314632

**Authors:** Myriam Fontaine, Kayla Horowitz, Nancy Anoja, Angela Genge, Kristiana Salmon

## Abstract

**Objective:** Genetic testing practices are rapidly evolving for people living with, or at-risk for, amyotrophic lateral sclerosis (ALS), due to emerging genotype-driven therapies. This study explored how individuals at-risk for familial ALS (fALS) perceive the opportunity to participate in a clinical trial, and to better understand how that may influence the decision-making process for predictive genetic testing.

**Methods:** This study used both quantitative and qualitative data analyses. Data were collected through an online questionnaire, followed by semi-structured interviews conducted with twelve (n=12) individuals at-risk for either *SOD1-* or *C9orf72*-ALS who had predictive testing prior to study participation. Interview data were analyzed using reflexive thematic analysis.

**Results:** Three overarching themes were conceptualized from the data: i) the psychosocial impact of fALS; ii) perspectives of at-risk individuals on research involvement; and iii) predictive genetic counselling and testing considerations. These results contribute perspectives of the lived experience to inform predictive genetic counselling and testing practices for individuals at-risk for fALS.

**Conclusion:** Individuals at-risk for fALS view potential participation in a presymptomatic clinical trial as an actionable measure that may increase their desire for predictive genetic testing. Genetic counselling was identified as a critical component of the predictive testing process given the life-changing implications associated with a positive result. Increased access to genetic counselling, and in a timely manner, is a significant need in the ALS population given potential access to gene-specific therapies in the presymptomatic stage.

## Introduction

Amyotrophic lateral sclerosis (ALS) is a progressive, adult-onset neurodegenerative condition, characterized by motor neuron degeneration, resulting in muscle weakness, eventual paralysis, and respiratory failure (1, 2). There is currently no cure. Underlying causes of ALS are still poorly understood in most cases; however, more than 40 genes have been associated with causing, or increasing risk of, ALS (1). While only 5-10% of people living with ALS have a family history of ALS or related conditions, such as frontotemporal degeneration (FTD), more than 15% carry a genetic variant associated with the disease. As a result, the historical distinction between ‘familial’ and ‘sporadic’ ALS has become less pertinent (3–5).

Predictive genetic testing is the process whereby asymptomatic individuals at-risk for a hereditary condition pursue genetic testing to determine if they carry a disease-causing genetic variant. Motivations for pursuing predictive testing have been well studied in other neurodegenerative conditions, such as Huntington’s Disease (HD) (6). Previous reports of perceived benefits include future planning, reproductive planning, relief of uncertainty and anxiety, and risk for children (7–13). Limited studies in ALS show similar results to those in HD and other neurodegenerative diseases (14–20). These studies also reveal reasons at-risk individuals do not pursue predictive testing, which include lived experience and trauma related to the condition, avoidance of guilt and anxiety, or simply a reluctance to know their status (7, 11, 12, 14–17, 21, 22). Often, at-risk individuals choose to not pursue predictive testing due to the perceived lack of treatment or preventative options.

The first gene-specific therapy is now approved for ALS caused by variants in *SOD1* (*SOD1*-ALS), and there is an ongoing clinical trial (NCT04856982), called ATLAS, for presymptomatic individuals who carry certain pathogenic variant in the *SOD1* gene (23). As such, predictive testing now has the potential to provide asymptomatic individuals with *SOD1* variants opportunities to participate in trials or gain access to genotype-driven therapies (13, 18, 22). In Canada, clinicians generally recognize clinical trials as sufficient to warrant the offer of genetic testing (24). However, this topic has not been thoroughly explored from the perspective of at-risk individuals.

There is a critical need for published guidelines on predictive genetic counselling and testing for ALS (25). This study explores how the prospect of clinical trials may influence the uptake of predictive testing for ALS and contributes to the development of any future guidelines.

## Methods

This study was conducted in accordance with the Tri-Council Policy Statement: Ethical Conduct for Research Involving Humans, and within requirements of the McGill University Health Centre Research Ethics Board (REB). Approval was obtained from the REB’s Neuroscience & Psychiatry Panel prior to study start (Project: 2023-8888).

### Sample

Participants had undergone predictive testing for a variant in either the *SOD1* or *C9orf72* genes within five years prior to study start. Participant inclusion focused on these autosomal dominant genes, as they are the most frequent causes of genetic ALS, and both have targeted therapies in development. Participants included some who pursued testing prior to initiation of the ATLAS presymptomatic *SOD1* trial (3/12), and some who pursued it after ATLAS began (9/12). During the study period, there were clinical trials targeting *C9orf72* expansions for symptomatic individuals; however, no presymptomatic, interventional *C9orf72-*ALS trials were ongoing or anticipated (though *C9orf72-*FTD trials were), in contrast to *SOD1*. Individuals with *C9orf72* repeat expansions are at-risk of symptoms beyond ALS, including FTD and parkinsonism; however, due to potential differences in lived experiences, participants had to have had a predominant family history of ALS to be included in the study.

Participants were recruited from across Canada, were at least 18 years old, had opted to have their test results disclosed to them, and spoke English or French. Those who declined disclosure of genetic results were excluded, though no potential participants met this criterion.

### Data Collection

This study utilized mixed-methods, including a questionnaire and semi-structured interviews based on recall. The questionnaire addressed four domains: 1) sociodemographic characteristics; 2) familial phenotype and lived experience with ALS; 3) predictive testing process; 4) factors influencing the decision to pursue testing. The interview guide was developed to elicit participants’ perceptions on clinical trials and their decision-making process for pursuing testing. Interviews were conducted in English or French by the bilingual first author, based on the participant’s preference. The interviewer kept a journal throughout the interview process to capture thoughts and reflections, to ensure reflexivity and acknowledge and address any biases related to her positionality as a healthy, non-disabled genetic counselling student (26, 27). All interviews were recorded through secure audio-visual conferencing and transcribed verbatim. Transcripts were de-identified prior to analysis. NVivo (Version R1.7.1) was utilized to organize data and code transcripts.

### Data Analysis

Quantitative data was analyzed using descriptive statistics. Qualitative data was analyzed using reflexive thematic analysis (26, 28) and narrative inquiry (29) to explore the nuanced experiences and decision-making processes of participants. These combined approaches uncovered complex perspectives, highlighting the multifaceted emotional, sociocultural, and ethical considerations influencing decisions within an evolving medical landscape.

The research team developed a thematic codebook using an inductive approach (28, 30), as two team members independently analyzed the first four transcripts to generate concepts of meaning and a codebook. Subsequently, the coding framework was refined through iterative and reflective discussion to reach a consensus. Remaining interviews were analyzed by the first author. This collaboration led to the generation of themes (31).

## Results

Twelve individuals participated in this study, with a summary of their characteristics in Table 1.

**Table 1.** Summary of participant characteristics.

|  | All participants<br>(n=12) | Participants at-risk for *SOD1*<br>(n=5) | Participants at-risk for *C9orf72*<br>(n=7) |
| --- | --- | --- | --- |
| Sex, female, *n* (%) | 6 (50.0) | 3 (60.0) | 3 (42.9) |
| Positive predictive testing result, *n* (%) | 8 (66.7) | 3 (60.0) | 5 (71.4) |
| Age, *n* (%) |  |  |  |
| 25-34 | 2 (16.7) | 1 (20.0) | 1 (14.3) |
| 35-44 | 3 (25.0) | 2 (40.0) | 1 (14.3) |
| 45-54 | 5 (41.7) | 2 (40.0) | 2 (28.6) |
| 55-64 | 1 (8.3) | 0 (0.0) | 2 (28.6) |
| 65-74 | 1 (8.3) | 0 (0.0) | 1 (14.3) |
| Province where predictive testing was received, *n* (%) |  |  |  |
| Alberta | 2 (16.7) | 2 (40.0) | 0 (0.0) |
| British Columbia | 2 (16.7) | 0 (0.0) | 2 (28.6) |
| Ontario | 1 (8.3) | 0 (0.0) | 1 (14.3) |
| Quebec | 7 (58.3) | 3 (60.0) | 4 (57.1) |
| Language of interview, *n* (%) |  |  |  |
| English | 9 (75.0) | 5 (100.0) | 4 (57.1) |
| French | 3 (25.0) | 0 (0.0) | 3 (42.9) |
| Ethnicity*, *n* (%) |  |  |  |
| White/Caucasian | 7 (58.3) | 5 (100.0) | 2 (28.6) |
| Native American | 1 (8.3) | 0 (0.0) | 1 (14.3) |
| French Canadian | 3 (25.0) | 0 (0.0) | 3 (42.9) |
| Sephardic Jewish | 1 (8.3) | 0 (0.0) | 1 (14.3) |
| Participants with children, *n* (%) | 6 (50.0) | 1 (20.0) | 5 (71.4) |

\*Options included in questionnaire for ethnicity mirror those used by Statistics Canada

The sample had equal representation of male and female sexes, through a range of ages. Participants predominantly self-identified as White/Caucasian and had a high degree of education: high school diploma (1/12), trade/technical/vocational training (1/12), college diploma (2/12), bachelor’s degree (5/12), and advanced degree (3/12). Half of the participants had biological children.

All participants lost a family member to ALS, two acted as caregivers for a relative with ALS (10/12 did not report a caregiver role), and five reported a family history of FTD. Participants underwent predictive testing between 2018 and 2023. Test outcomes consisted of eight positive results and four negative results. One participant reported experiencing symptoms of ALS at the time of the interview, while the remainder reported being asymptomatic.

### Quantitative Data

In the questionnaire, many participants (7/12) considered the opportunity to participate in a clinical trial as an actionable measure for individuals who receive a positive result. Additionally, participants ranked a series of common factors known to influence decision-making for predictive testing, as described in the previous literature. Factors were attributed a score, where a score of “1” was attributed to the factor ranked highest by the participant (most important), and “9” (motivating factors) or “10” (demotivating factors) was attributed to the factor ranked lowest (least important). The mean rank and standard deviation were calculated and are summarized in Tables 2 and 3, alongside the number of participants who selected each factor as one that influenced their decision-making. While the sample size is underpowered, ‘hope to be eligible for future treatments’ scored high as a motivating factor, and recurrently among participants (10/12). Conversely, the ‘lack of effective treatment/preventative measures available’ ranked high as a demotivating factor, but less recurrently (2/12).

**Table 2.** Potential motivators that contribute to decision-making for predictive genetic testing.

| Which factors motivated you to pursue predictive testing for ALS? (Choose all that apply) |  |  |  |
| --- | --- | --- | --- |
| Factor | # participants that selected this as having influenced their decision-making (n=12) | Mean ranking | Standard Deviation |
| Relieve uncertainty/reduce anxiety | 9 | 3.42 | 2.07 |
| Hope to be eligible for future treatments as research advances | 10 | 3.50 | 1.45 |
| Plan for the future (career, finances, priorities) | 10 | 3.92 | 2.47 |
| Chance to participate in a clinical trial for presymptomatic individuals | 5 | 4.42 | 1.78 |
| Family history and/or personal experience caring for a relative with ALS | 6 | 4.83 | 2.89 |
| Provide information to relatives | 4 | 6.08 | 1.73 |
| Confirm my gut feeling | 2 | 6.17 | 2.62 |
| Inform the risk for my children | 4 | 6.33 | 2.84 |
| Family planning/reproductive decision-making | 4 | 6.33 | 3.17 |

**Table 3.** Potential deterrents that contribute to decision-making for predictive genetic testing.

| Which factors were not motivating in the pursuit of predictive testing for ALS? (Choose all that apply) |  |  |  |
| --- | --- | --- | --- |
| Factor | # participants that selected this as having influenced their decision-making (n=12) | Mean ranking | Standard Deviation |
| Worry or anxiety about coping with results | 1 | 4.25 | 2.73 |
| Lack of effective treatment/preventative measures available | 2 | 4.33 | 3.17 |
| Personal experience and/or traumatic memories associated with ALS | 2 | 4.83 | 3.04 |
| Worry about insurance and/or discrimination | 4 | 4.83 | 3.69 |
| Did not want to upset my family and friends with my result | 1 | 5.00 | 2.00 |
| Feelings of guilt for passing the familial mutation onto children | 1 | 5.67 | 1.97 |
| Lack of access to clinical trials/research opportunities | 1 | 5.75 | 2.22 |
| Fear or survivor guilt if found not to carry the familial mutation | 1 | 6.25 | 3.08 |
| Not wanting to know my carrier status/not ready to hear the information | 1 | 6.75 | 2.30 |
| Barriers in accessing genetic testing services (i.e. long waitlist) | 3 | 7.33 | 3.39 |
| None of the above |  |  |  |

### Qualitative Analysis

Overarching themes from participant interviews are summarized, with a selection of supporting quotes, in Table 4.

**Table 4.** Summary of reflexive thematic analysis.

|  |
| --- |
| **THEME 1: THE PSYCHOSOCIAL IMPACT OF GENETIC ALS** |
| **Sub-Theme 1: Adaptation and coping mechanisms** |
| **A. Overwhelming negative perception of ALS:** *It's the most horrible thing you could ever have happened to you, ... it's relentless in taking things away and it takes everything away. (001-EN)* |
| **B. Reactions to a familial diagnosis:** *I can tell you when I found out the results of [my aunt's] test and then connecting it with my cousin's test, it hit me like a brick wall. I had panic attacks; I was devastated because then I knew it was in our family. I knew I had a 50 percent chance. (004-EN)* |
| **C. Mental preparation prior to genetic testing:** *I had convinced myself that I was positive. ... I think naturally I expect the worst, and I already do some of the grieving, ... so that when [I get bad news], it's a little less worse. (007-FR)* |
| **D. Acceptance of personal result:** *For whatever reason, I kind of was prepared for it. So when I did get the news, I'm not going to say I was stoic, but it was just a matter of fact. ... There's still that one brief exhale where you're like, okay, yeah, rats, ... you always hold out a little bit of hope. (011-EN)* |
| **E. Positive reappraisal:** *I just have the outlook of 'live life to the fullest'. So yes, I know I have it, but do I want to think about it every day and bring everybody that loves me and is with me right now down? No. So trying to be as positive as I can. (010-EN)* |
| **Sub-Theme 2: Impact on relationships** |
| **F. Experiencing the disease through an affected family member:** *Those are difficult things when you're cleaning up a man who you've looked at and idolized your whole life. ... Those are things that I had trouble reconciling. I don't regret being there for him, but I do suffer with those memories a little bit. (011-EN)* |
| **G. Bonding with other family members:** *We like bonded together in this crappy news. It's like we're two castaways and we're sharing a buoy, and we're happy to be sharing the same buoy even if both of us are going to drown ... [at least] we have the buoy together. (005-FR)* |
| **H. Strain with other family members:** *I feel like my relationships with my siblings have been affected. And I feel like I'm not let in the same way. And I have an extreme sense of guilt that I didn't know would be there. ... It's overwhelming. (012-EN)* |
| **I. Communicating risk to children:** *I have not told either of my children for a variety of reasons that I have tested and that my C9 is mutated. I have not shared that with them, just about I can't. (003-EN)* |
| **THEME 2: PERSPECTIVES ON RESEARCH INVOLVEMENT** |
| **Sub-Theme 1: Benefits** |
| **A. Actionable:** *I think that fundamentally, when there is nothing else, clinical trials represent ... some modicum of treatment. (006-EN)* |
| **B. Altruism:** *Regardless, science is bigger than every individual. ... I consider that science is the real important thing ... to advance research ... progress for the common good. (005-FR)* |
| **C. Hope:** *I think there's a real reason for people to have hope. There's a real reason now, where say 10 years ago, there wasn't. (001-EN)* |
| **Sub-Theme 2: Apprehensions towards research involvement** |
| **D. Wariness of effectiveness:** *I would sit back, ... until [I] find out, is this going to work, is it actually effective? And you have to be tested to go into that trial. I would wait until it's more of a sure thing. (004-EN)* |
| **E. Potential negative impacts:** *Let's not talk about just the obvious cons that this drug might not work, or it may have negative effects. Let's talk about all the negative effects. ... Because you don't want to have somebody in a trial that never asked you these questions ... and all of a sudden, they're having some severe financial or emotional or mental problems. (001-EN)* |
| **F. Desire to resist hope:** *[I wanted] just to be able to accept it, this disease, ... For it not to be a race, my life, for a treatment ... that for me, it's already a fatality that I will die from it. (007-FR)* |
| **Sub-Theme 3: Perspectives on trial design** |
| **G. Understanding the need for placebos:** *I understand how that works. I mean obviously, I would be praying to have the drug ... [but] it's part of how you make a trial legitimate, right? It's necessary. (012-EN)* |
| **H. Clinical trial design considerations for the placebo:** *I would hope, I would anticipate that the placebo group would get some kind of off label advanced access to the drug were it to be a positive clinical trial. So, it's not I get it, or I never get it, it'll be I get it, or I get it a little bit later if it works. (006-EN)* |
| **I. Genetic testing eligibility criterion is inevitable:** *If we want targeted therapies, we kind of need to narrow down our study population ... or else we're just diluting the effects. (006-EN)* |
| **THEME 3: PREDICTIVE TESTING CONSIDERATIONS** |
| **Sub-Theme 1: Primary motivators remain conventional** |
| **A. Relieve uncertainty:** *I don't like that ominous black cloud that could be or maybe won't be, ... I'd much rather know with certainty if I'm going to potentially deal with something, or at least that it's in my reality. (011-EN)* |
| **B. Family planning:** *We'd like to have more children, so being able to prevent the transmission of this gene, it's also something that interested us because I know it's possible. (005-FR)* |
| **C. Inform risk for family members:** *As far as the testing, ... a good part of it was, in the event I was negative, ... I wanted to assure my kids they were out of the woods because it has an impact for life insurance and career decisions and family decisions and all sorts of things. (003-EN)* |
| **Sub-Theme 2: Influence of emerging therapies** |
| **D. Enhancer but not primary motivator:** *I already knew I was going to do it. ... It just tipped the scale for me. It was just sort of a no brainer. (012-EN)* |
| **E. Provides hope:** *I feel that knowing that [I]'d have [a clinical trial] as something [I] could partake in [if I test positive] would be valuable for me to know in advance. ... I had enough boxes ticked that I wanted to get the test done anyways, but had I known ... that would definitely make a difference for me for sure because it gives some hope. (012-EN)* |
| **F. Lack of perceived urgency in absence of clinical trials:** *At the time, I kind of did a quick scan ... and I didn't necessarily see a genetic test result of C9orf as anything especially actionable at the time. It didn't really seem like information that I could do very much with. (006-EN)* |
| **Sub-Theme 3: Predictive testing is a highly individual decision** |
| **G. Differing stances within families:** *No, no testing [for my daughter]. I actually don't even know if she realizes that it's a 50 percent chance. ... But I can't control that, and I'm not going to ever ... push anything on her, so I just put that in a box, store it and leave it. (004-EN)* |
| **H. Individual perceived barriers:** *I don't want my children prejudged. I don't want the insurance companies canceling [my children] out because they have a potentially mutated gene. (003-EN)* |
| **I. Context of current life stage:** *Unless you're really doing some sort of family planning or financial planning, there's no reason to really know ... except that you're going to end up with this horrendous disease at the end. (012-EN)* |
| **Sub-Theme 4: Importance of genetic counselling** |
| **J. Impact of genetic testing is bigger than science:** *We're playing with death, ... or the prediction of death. ... It's not simple. I think you need to be solid between both ears to take this on. If you're not sure, look, maybe you shouldn't play with that. (008-FR)* |
| **K. Pre-test counselling needs:** *I think that before getting tested, it's more than a 15-minute meeting with the doctor. ... This is a life changing result. ... [People] need to really understand what they're in for, what the results could mean for them, and what the results may or may not give them access to. (006-EN)* |
| **L. Genetic counselling in the context of clinical trials:** *There's too many factors, there's too many what ifs in the clinical trial. And I think that all of those need to be made clear at the moment of genetic counseling, long before we stick a needle into anyone. ... There's so many implications [to] this, and I would hate to think that people are making that decision just based on maybe I'll get into a trial. (006-EN)* |

### Theme 1 – Psychosocial Impact of fALS

#### Adaptation and coping mechanisms

Participants had an overwhelmingly negative perception of ALS, describing it as a ‘*horrific*’ (001-EN; 012-EN) disease with a physical, mental, and emotional toll. Feelings of distress, grief, and bereavement were triggered as participants came to the realization that they themselves were at-risk. When evaluating the potential of a positive result, participants described anticipatory grief, using an ‘*expecting the worst*’ approach, whether intentional or not, as ‘*a defensive mechanism*’ (006-EN):

> *‘I had convinced myself that I was positive … I think naturally I expect the worst, and I already do some of the grieving … so that when [I get bad news], [I hope] it’s a little less worse’ (007-FR)*

Participants who tested positive adapted differently, some struggling with the intangibility of being a genetic carrier, and others accepting this as ‘*just a matter of fact*’ (011-EN).

#### Impact on relationships

For many, the sentiment of a shared experience facilitated communication within the family and created a built-in support network:

> *‘We bonded together in this crappy news. It’s like we’re two castaways and we’re sharing a buoy, and we’re happy to be sharing the same buoy even if both of us are going to drown … [at least] we have the buoy together.’* (005-FR)

Contrarily, some mentioned this is not the case for all relatives and can cause strain on existing relationships. Participants experienced pre-emptive feelings of guilt at the time of testing with regards to their children. Participants struggled with risk communication to their children, expressing ‘*[it’s] the hardest part*’ (011-EN).

### Theme 2 – Perspectives on research involvement

#### Benefits

Most participants saw clinical trials as something actionable that people at-risk for ALS can pursue, expressing a sentiment of ‘*why would you not [participate]’* (009) if given an opportunity:

> ‘*I think that fundamentally, when there is nothing else, clinical trials represent … some modicum of treatment.*’ (006-EN)

Most emphasized that simply getting involved in research to ‘*hopefully be part of the cure one day*’ (002-EN) was perhaps even more important than receiving an experimental therapy. Participants spoke about their ‘*general optimism*’ (006-EN) regarding future therapies and highlighted that, with emerging clinical trials, ‘*there’s a real reason for people to have hope*’ (001-EN).

#### Apprehensions towards research involvement

Questions arose around trial participation requirements and potential outcomes associated with their participation, given that the effectiveness of an experimental therapy is unknown. Some voiced concerns regarding potential downsides, such as time away from family, financial burden, health risks, and added psychological stress. Others considered the potential downsides as insignificant, stating ‘*who cares*’ (009-EN), given the potential future benefit.

#### Perspectives on trial design

All participants expressed an acceptance for use of placebo groups in clinical trials:

> ***‘****I understand how that works. I mean obviously, I would be praying to have the drug* (*20*) *… [but] it’s part of how you make a trial legitimate, right? It’s necessary.’ (012-EN)*

Many voiced a hope that trial design would allow for early drug access for the placebo group, with one stating this as a non-negotiable: ‘*that’s the only way I’d do it*’ (004-EN). However, most concluded that, in the grand scheme of things, ‘*no, [the placebo] wouldn’t stop me*’ (005-FR) and they would still ‘*roll the dice*’ (009-EN). Participants also agreed that it is *‘logical’* (005-FR) for genetic testing to be mandatory for inclusion.

### Theme 3 – Predictive testing considerations

#### Primary motivators remain conventional

Regarding motivations for pursuing predictive testing, many emphasized the need to relieve uncertainty as ‘*undeniable*’ (007-FR) and the drive to ‘*end that war one way or the other*’ (004-EN). The majority discussed clarifying their genetic carrier status as a means of ‘*organizing life*’ (007-FR), and concretizing life plans in the event of a positive result:

> *‘I wanted to go get to the bottom of it because … if I’m positive, the plan was … that we would financially secure my wife’s future and then with what would remain … we would retire together for as long as it lasted.’* (008-FR)

This same motivation was brought forth by those currently in the family planning stage. For most participants with children, informing future generations of risk was important.

#### Influence of emerging therapies

Many described clinical trials as ‘*an added bonus’* (010-EN), but not their primary motivator. All five participants from the *SOD1* group were aware of potential upcoming trial opportunities, and ‘*figured by knowing, there would be opportunities [to be part of research]*’ (011-EN).

> ‘*I just wanted to get on it in case I was positive, and I could be a part of the trials*.’ (002-EN)

Comparatively, a lack of perceived urgency was described by participants for whom presymptomatic trials were not yet available (i.e. *C9orf72*). However, these same participants hypothesized that presymptomatic trials may become a stronger motivator in the future, but only once they are a reality.

#### Predictive testing is a highly individual decision

Stances on predictive testing varied within participants’ families, highlighting that it’s ‘*a very personal decision*’ (006-EN). Each participant’s personal life influenced the decision to test and the timing. When exploring what might decrease motivation to pursue testing, insurance and fear of discrimination were perceived barriers:

> ‘*I don’t want the insurance companies canceling [my children] out because they have a potentially mutated gene*.’ (003-EN)

#### Importance of genetic counselling

Participants emphasized a need for comprehensive genetic counselling to discuss the implications of learning one’s status, and what this may represent in the context of therapeutic access:

> ‘*Before getting tested, it’s more than a 15-minute meeting with the doctor. … This is a life-changing result. [People] need to really understand what they’re in for, what the results could mean for them, and what the results may or may not give them access to*.’ (006-EN)

## Discussion

Participants re-affirmed that a diagnosis of fALS impacts relationships with one’s relatives and children. Building upon previous research (32), the bond formed through shared experience was highlighted, as many expressed increased feelings of closeness with at-risk relatives. Still, participants noted that, with discordant coping mechanisms, a diagnosis of fALS may disrupt communication and strain relationships, especially with relatives who aren’t as acknowledging of the disease and associated risk, which aligns with previous research. Similarly to other inherited genetic conditions, risk communication to children remains a challenge and an emotional burden.

Clinical trial participation is seen as a significant, actionable opportunity for those with positive genetic test results. This aligns with the perspectives of Canadian clinicians (24) who perceive the opportunity to participate in a trial as an actionable treatment plan. The relationship between research involvement and hope was a major theme elucidated in this study. This included the hope of directly benefiting from an experimental drug, contributing to research that has the potential to lead to a therapy, or both. Altruism as a coping mechanism was clearly conveyed. These findings support the importance of integrating discussions of research opportunities, and possibly referrals to other centres, in predictive genetic counselling discussions. Additionally, those at-risk for genetic ALS/FTD are calling upon the medical and scientific community for earlier access to intervention options (33), and many study participants expressed a desire to support advocacy efforts, specifically with regards to treatment and care of at-risk individuals.

Placebos were a well-understood concept, and participants had an overall neutral opinion regarding their use in clinical trials. However, participants were uncomfortable with the thought that the placebo group may not have access to the experimental therapy, if therapeutic effect was demonstrated during the placebo-controlled portion. To counter this perceived barrier, participants explored the possibility of having immediate initiation of the experimental therapy at the first sign of therapeutic effect or symptoms. This design concept was brought forth organically by participants, without any prompting from the interviewer. Some participants expressed concern regarding timely access to predictive testing, and suggested an integration of genetic counselling and testing into the trial protocol as a solution. The ATLAS study addresses these concerns, and therefore its design could serve as a model for future presymptomatic trials (23).

These findings identify conventional factors such as relieving uncertainty, future planning, and informing risk as remaining the primary motivators for at-risk individuals(13–20). Therefore, those providing predictive genetic counselling to this population can feel comfortable continuing to practice within conventional considerations (6).

Participants elucidated apprehensions that reduced their motivation to pursue predictive testing, including insurance concerns and fear of discrimination. Participant narratives further support the importance of discussing insurance before pursuing predictive testing and helping individuals navigate the insurance system, where possible (34). The Genetic Non-Discrimination Act was recently upheld at the Supreme Court of Canada (35); however, it remains to be seen how effective it is in practice.

Finally, genetic counselling was highlighted as critical. Participants described a need to better understand all potential life implications associated with any result (positive, negative, or uncertain), a need for thorough self-reflection and mental preparation, and a need for increased access to research advances, especially pertaining to research opportunities. While guidelines have been published for those diagnosed with ALS (36), consensus on predictive testing is lacking (25). Canadian clinicians have indicated they prefer to refer at-risk individuals to a genetics service (24), further supported by research indicating neurologists have low genomic knowledge and counselling skills compared to genetics specialists (37). However, long waitlists and a lack of genetic counselling services threaten to compromise quality of care, especially outside of metropolitan areas and the global North. Given the emerging gene-targeted therapies that will be available to a subset of this population, it is essential to not only have standardized predictive genetic counselling and testing guidelines informed by lived experience, but to ensure the proper infrastructure is in place to provide these services in a timely manner.

### Limitations

This study is limited by its small sample size and restricted geography, limiting transferability to developed nations of, primarily, European descent. Other limitations include recall bias and memory decay. Results may not be generalizable to other ALS variants, or variants with distinct phenotypes (e.g. *FUS*-ALS with juvenile onset). Participants with a family history of solely FTD were excluded, and therefore their experiences are not represented in this work and further research is needed.

## Conclusion

These findings enrich the scope of predictive genetic counselling considerations for ALS, offering insights for the development of future practice guidelines. The study demonstrated that traditional motivators still drive predictive testing, despite new therapeutic developments. It also underscores the importance of better access to research information, especially with the advent of gene-targeted therapies, to enhance decision-making. Genetic counsellors and other healthcare providers, as appropriate, should consider introducing discussions about ongoing ALS research in pre-test counselling, and adapt to individual informational needs. These findings provide a glimpse into the potential weight that interventional presymptomatic clinical trials may hold for future decision-making, how predictive testing uptake may increase, and a need for increased genetic counselling services as a result.

## Acknowledgments

The authors thank the study participants, who intimately shared their personal journeys to help enhance the experience for those who will explore predictive testing in the future. The authors also thank Laynie Dratch and Jean Swidler for their review and feedback on this manuscript.

## Data availability

Anonymized, collated data may be provided by the corresponding author (KS) upon reasonable request and appropriate permissions.

## Author Contributions

MF was responsible for development of the research question, literature review, questionnaire development, interview guide construction, REB submission, interview conduct, data analysis, and manuscript writing and editing. KH acted as second coder for qualitative data analysis, and provided manuscript writing and editing. NA contributed to the project conceptualization and study design. AG contributed to the REB submission and provided medical oversight. KS contributed to the project conceptualization, study design, REB submission, participant recruitment, data analysis, manuscript writing and editing, and project oversight.

